# Sex Differences in the Protective Effect of Brain Volume: Age Attenuates Protection in Females

**DOI:** 10.1101/2025.10.30.25339193

**Authors:** Javier Gomez-Farias, Ngoc Mai Le, Joseph N. Samaha, Bruna Kfoury, Hussain M. Azeem, Ritesh Bajaj, Shikha Tripathi, Shayan Shams, Luca Giancardo, Eunyoung Lee, Robert Regenhardt, Louise D. McCullough, Sunil A. Sheth

## Abstract

**Background:** Sex differences in outcomes after acute ischemic stroke (AIS) are well recognized, but mechanisms remain unclear. This study evaluates whether parenchymal brain volume (PBV), age, and final infarct volume (DWI-FIV) explain sex-specific differences in post-stroke disability.

**Methods:** We analyzed a prospectively collected multicenter registry of AIS patients treated between 2015 and 2024. Automated pipelines quantified PBV from non-contrast CT and DWI-FIV from 24–48-hour MRI. With the primary outcome being 90-day functional independence (mRS 0–2), sex-stratified logistic regression models evaluated its association with PBV. Logistic and linear models examined sex-specific relationships among covariables.

**Results:** Among 1,103 patients, 48% were female, median age was 66 [IQR 56–77], median NIHSS was 9 [IQR 4-17], and median DWI-FIV was 7.8 [IQR 0.2-36.3]. Functional independence at 90 days was achieved in 51.7% of males versus 44.9% of females (*p* = 0.025). In univariable analyses, greater PBV was associated with higher odds of functional independence in both males (OR 1.30; 95% CI: 1.11–1.53) and females (OR 1.35; 95% CI: 1.13–1.61). After ruling out multicollinearity (mean VIF = 1.32), multivariable analyses showed persistent association in males (aOR 1.30 per 100 cm^3^; 95% CI: 1.10–1.60) but not in females (aOR 1.10; 95% CI: 0.90–1.33). Each decade of age reduced the odds of functional independence by 30% in females (aOR 0.70; 95% CI: 0.62– 0.79) versus 15% in males (aOR 0.85; 95% CI: 0.72–0.97; *p i*nteraction = 0.019). Age was also associated with larger infarcts in females (+4.22 cm^3^/decade; *p* = 0.033) but not in males.

**Conclusion:** PBV was protective in males but less so in females, where stronger age-related effects on infarct size and severity abrogated its benefit. These suggest aging weakens brain reserve in women, emphasizing the importance of sex and brain volume measurements in outcome models.

## Introduction

Acute ischemic stroke (AIS) outcomes differ by sex, with females generally experiencing greater initial stroke severity, higher rates of disability, and less functional independence compared to men, yet the mechanisms underlying these differences remain unclear.^1^ ^2^ ^3^ ^4^ Data on sex-based outcome disparities are inconsistent; some studies demonstrate clear differences in post-stroke disability and mortality, while others show no significant differences after adjusting for age, stroke severity, and infarct size. ^3^ ^4^ ^5^ ^6^ The clinical literature further demonstrates that females are typically older at stroke onset, have a higher prevalence of atrial fibrillation, and are more likely to present with pre-existing disability, all of which may contribute to poorer outcomes. ^1^ ^7^ ^8^ ^9^ However, the relationship between these variables and sex-specific outcomes remains incompletely understood. ^4^

Parenchymal brain volume (PBV) may be an important and underappreciated contributor to sex differences in AIS outcomes. PBV reflects the total amount of functional brain tissue available, which can influence the clinical impact of a given infarct. ^10^ ^1^ ^11^ ^4^ Females, who tend to be older at stroke onset and have smaller absolute brain volumes, may have greater white matter disease burden as a similar lesion may represent a larger proportion of total brain tissue lost. For this reason, the protective effect of PBV may diminish with age and comorbidities. ^12^ ^13^ ^14^

In addition, final infarct volume (FIV) is another independently established predictor of post-stroke outcomes; larger infarcts are associated with worse functional recovery and greater long-term disability. ^14^ ^15^ ^16^ However, the impact of FIV may be modulated by PBV and age, and these interactions could contribute to observed sex differences in recovery trajectories. In this study, we analyzed a large multicenter cohort using automated neuroimaging segmentation to quantify infarct volume and total parenchymal brain volume and determine how these features interact and contribute to sex differences in outcomes after AIS.

## Methods

### Data Availability

The data used in this study are available for collaborative groups, provided there is approval from the institutional review boards and data-sharing agreements in place. This study followed the Strengthening the Reporting of Observational Studies in Epidemiology (STROBE) reporting guideline, and the study protocol was reviewed and approved by the UTHealth Institutional Review Board.

### Participants

From our prospectively collected multicenter registry from 4 comprehensive stroke centers and 5 primary stroke centers, we identified patients with AIS who were treated between February 1, 2015, and March 31, 2024, in the Greater Houston area. We included patients with a documented 90-day mRS score. Patients were excluded if they had registration or segmentation errors or if they had incomplete data for outcomes or predictors (n=276).

### Measurements

PBV was determined using an automated pipeline applied to the brain CT. The pipeline begins with skull-stripping the raw brain CTs using SynthStrip ^17^, a brain extraction tool with the default value of 1 mm for the mask border threshold. The skull-stripped CTs are registered to a standard MNI152T1 template using FSL version 5 with FLIRT, ^18^ a linear registration tool.

An automated threshold selection technique, Otsu’s method, ^19^ was employed to segment the parenchymal blood volume from the registered brain CT images. Prior to thresholding, a brain mask was generated by applying an intensity threshold of –300 Hounsfield Units (HU) to the CT volume, isolating the brain region from surrounding structures. The masked image was then intensity-clipped to the range [–15, 80] HU to retain voxel values most representative of brain parenchyma.

Otsu’s method was subsequently applied to the clipped image within a restricted intensity range of [–5, 30] HU to compute a global threshold that optimally separates brain parenchyma from non-parenchymal regions. This threshold was applied to the clipped volume, and the result was further refined using the mask, yielding the parenchymal segmentation. The total parenchymal volume was quantified by multiplying the number of voxels within parenchymal segmentation by the unit voxel volume (in mm^3^), with the final measurement expressed in cubic centimeters (cm^3^). FIV was defined using an automated core volume segmentation pipeline, using the diffusion-weighted imaging (DWI) and ADC MRI obtained 24-48 hours after stroke. The pipeline uses nnU-Net version 2 to obtain the segmentation using the 3D DWI and ADC MRI. nnU-Net ^20^ ^21^ is a self-configuring deep learning based on automated semantic segmentation methods. The DWI-FIV was calculated from the total number of voxels belonging to the segmented core multiplied by the unit voxel volume (in mm^3^), and the final measurement was expressed in cm^3^.

### Outcomes

The primary outcome was functional independence at 90 days, defined as a modified Rankin Scale (mRS) score of 0–2. The primary exposure of interest was parenchymal brain volume (PBV), scaled to 100 cm^3^ and was assessed using multivariable regressions as detailed in the section below.

### Statistical Analysis

Descriptive statistics were presented as mean and standard deviation for continuous variables with a normal distribution and as median with interquartile range for those with a non-normal distribution. Categorical variables were summarized using absolute frequencies and proportions. Baseline characteristics were compared between sexes using the Wilcoxon rank-sum test for continuous variables and the Chi-square test for categorical variables.

For the primary analysis, sex-stratified univariable and multivariable logistic regression models were used to evaluate the association between PBV (per 100 cm^3^) and functional independence at 90 days. Multivariable models were adjusted for potential confounders, including final infarct volume (DWI-FIV), age, and baseline NIHSS. Model estimates were reported as odds ratios (ORs) with 95% confidence intervals (CIs), and statistical significance was defined as a two-sided *p* value < 0.05.

For the secondary analyses, we included pre-stroke mRS, as this variable could potentially bias the direction and magnitude of the PBV effect. We performed targeted analysis on the role of pre-stroke mRS on sex differences in outcomes. The proportion of missing data on pre-stroke mRS was assessed to guide the handling strategy. Multiple imputation (MI) was implemented to enhance statistical efficiency and model robustness for this analysis. Complete-case analysis (CCA) results were compared with the imputed analyses to evaluate the consistency of findings.

Logistic regression models were used to examine the associations of age, NIHSS, and DWI-FIV with functional independence at 90 days. Each model incorporated a sex-by-variable interaction term to assess whether these relationships differed between males and females.

To further explore the interrelationships among covariates, we conducted a series of pairwise linear regression analyses. These models assessed the bivariate associations between age, NIHSS, and DWI-FIV, each modeled as a dependent–independent pair (e.g., age vs. DWI-FIV, age vs. NIHSS, NIHSS vs. DWI-FIV). The primary pairwise models assessing the relationship between age and DWI-FIV, stratified by sex, are presented in the main text. Additional exploratory sex-stratified models evaluating associations between age and NIHSS, and between NIHSS and FIV, are provided in the Supplementary Methods and Figures S1–S2.By examining these relationships separately for each sex, we aimed to characterize how shared variance among these covariates may influence PBV’s association with functional independence at 90 days. Marginal effects were estimated to quantify sex-specific associations.

### Sensitivity Analyses

Sensitivity analyses replicated the primary multivariable models with 90-day mRS functional independence as an outcome while restricting the cohort to patients without large vessel occlusion (non-LVO) to assess the robustness of the findings.

Statistical significance was defined as a two-sided α level of 0.05, and all analyses were performed using Stata version 18.5 (StataCorp LLC, College Station, TX).

## Results

Among 1,103 patients with AIS, the median age was 66 years [IQR, 56-77], median NIHSS score was 9 [IQR, 4-17], median PBV was 1,698 cm^3^ [IQR, 1,611-1,759], median ASPECTS was 10 [IQR, 8-10], and median CTP-predicted infarct core volume was 3.0 cm^3^ [IQR, 0.0-21.0] (Table 1). The cohort included 528 females (48%) and 575 males (52%). Females were significantly older, had greater initial NIHSS, and larger CTP-predicted infarct core volumes compared with males (Table 1). No significant sex differences were observed in pre-mRS, DWI-FIV, PBV, and past medical history, including atrial fibrillation, hypertension, prior stroke, and other comorbidities (Table 1). In the univariate analysis, males had a significantly greater rate of functional independence at 90-day (51.7%, n= 297) compared with females (44.9%, n=237; *p* = 0.025). Mortality was significantly greater among females (24.1%, n=127) than males (17%, n=98; *p* = 0.004) in the univariate analysis. There were no significant sex differences in the rates of excellent functional independence at 90-days (*p* = 0.081) in the univariate analysis.

**Table 1.**
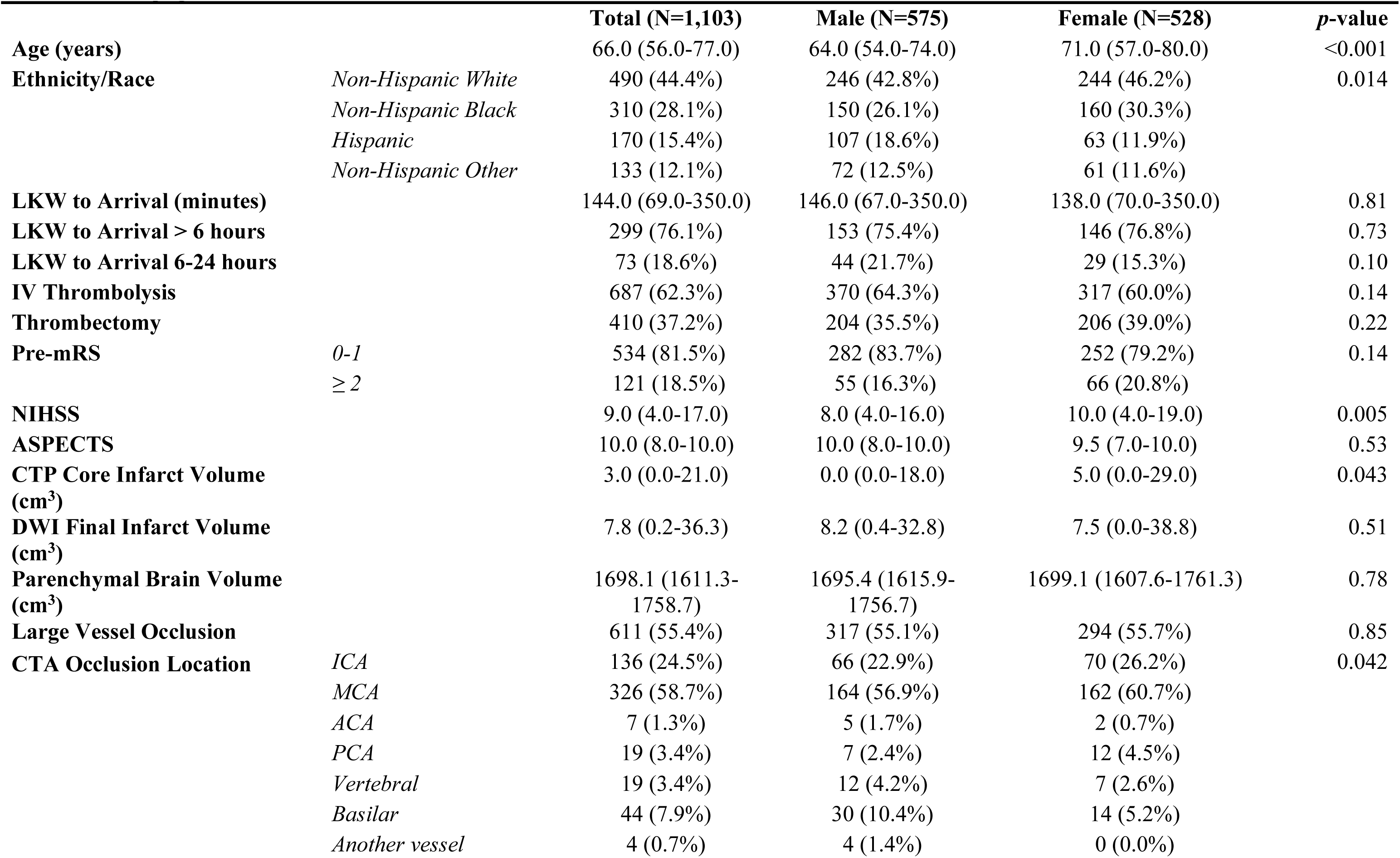
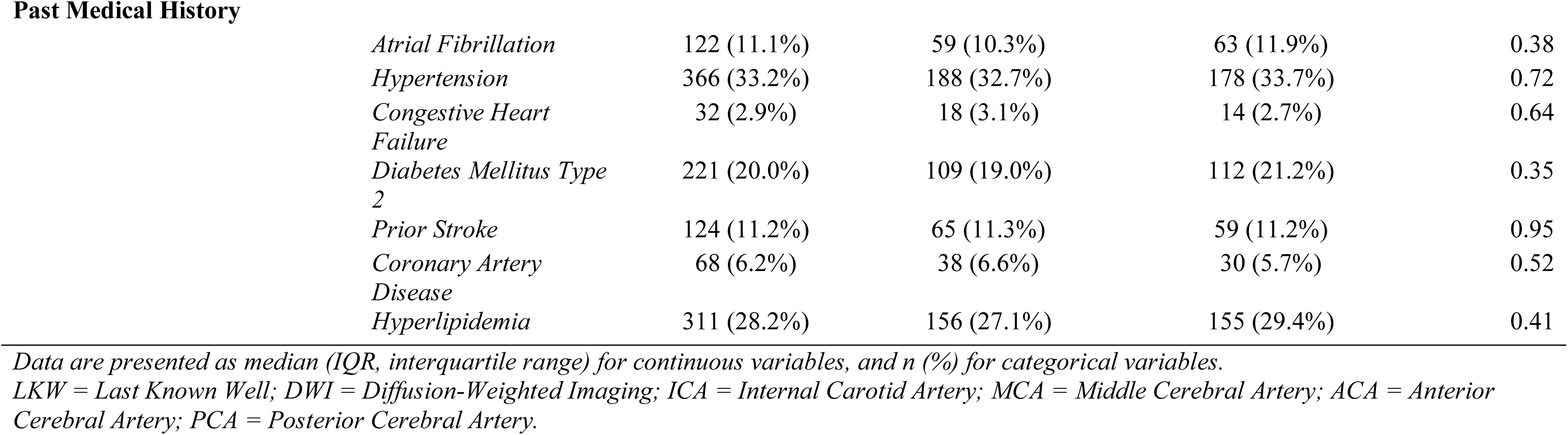
Demographics, baseline characteristics, and comorbidities.

|  |  | **Total (N=1,103)** | **Male (N=575)** | **Female (N=528)** | ***p*-value** |
| --- | --- | --- | --- | --- | --- |
| **Age (years)** |  | 66.0 (56.0-77.0) | 64.0 (54.0-74.0) | 71.0 (57.0-80.0) | <0.001 |
| **Ethnicity/Race** | *Non-Hispanic White* | 490 (44.4%) | 246 (42.8%) | 244 (46.2%) | 0.014 |
|  | *Non-Hispanic Black* | 310 (28.1%) | 150 (26.1%) | 160 (30.3%) |  |
|  | *Hispanic* | 170 (15.4%) | 107 (18.6%) | 63 (11.9%) |  |
|  | *Non-Hispanic Other* | 133 (12.1%) | 72 (12.5%) | 61 (11.6%) |  |
| **LKW to Arrival (minutes)** |  | 144.0 (69.0-350.0) | 146.0 (67.0-350.0) | 138.0 (70.0-350.0) | 0.81 |
| **LKW to Arrival > 6 hours** |  | 299 (76.1%) | 153 (75.4%) | 146 (76.8%) | 0.73 |
| **LKW to Arrival 6-24 hours** |  | 73 (18.6%) | 44 (21.7%) | 29 (15.3%) | 0.10 |
| **IV Thrombolysis** |  | 687 (62.3%) | 370 (64.3%) | 317 (60.0%) | 0.14 |
| **Thrombectomy** |  | 410 (37.2%) | 204 (35.5%) | 206 (39.0%) | 0.22 |
| **Pre-mRS** | *0-1* | 534 (81.5%) | 282 (83.7%) | 252 (79.2%) | 0.14 |
|  | *≥ 2* | 121 (18.5%) | 55 (16.3%) | 66 (20.8%) |  |
| **NIHSS** |  | 9.0 (4.0-17.0) | 8.0 (4.0-16.0) | 10.0 (4.0-19.0) | 0.005 |
| **ASPECTS** |  | 10.0 (8.0-10.0) | 10.0 (8.0-10.0) | 9.5 (7.0-10.0) | 0.53 |
| **CTP Core Infarct Volume (cm<sup>3</sup>)** |  | 3.0 (0.0-21.0) | 0.0 (0.0-18.0) | 5.0 (0.0-29.0) | 0.043 |
| **DWI Final Infarct Volume (cm<sup>3</sup>)** |  | 7.8 (0.2-36.3) | 8.2 (0.4-32.8) | 7.5 (0.0-38.8) | 0.51 |
| **Parenchymal Brain Volume (cm<sup>3</sup>)** |  | 1698.1 (1611.3-1758.7) | 1695.4 (1615.9-1756.7) | 1699.1 (1607.6-1761.3) | 0.78 |
| **Large Vessel Occlusion** |  | 611 (55.4%) | 317 (55.1%) | 294 (55.7%) | 0.85 |
| **CTA Occlusion Location** | *ICA* | 136 (24.5%) | 66 (22.9%) | 70 (26.2%) | 0.042 |
|  | *MCA* | 326 (58.7%) | 164 (56.9%) | 162 (60.7%) |  |
|  | *ACA* | 7 (1.3%) | 5 (1.7%) | 2 (0.7%) |  |
|  | *PCA* | 19 (3.4%) | 7 (2.4%) | 12 (4.5%) |  |
|  | *Vertebral* | 19 (3.4%) | 12 (4.2%) | 7 (2.6%) |  |
|  | *Basilar* | 44 (7.9%) | 30 (10.4%) | 14 (5.2%) |  |
|  | *Another vessel* | 4 (0.7%) | 4 (1.4%) | 0 (0.0%) |  |

Past Medical History
|  |  |  |  |  |
| --- | --- | --- | --- | --- |
| *Atrial Fibrillation* | 122 (11.1%) | 59 (10.3%) | 63 (11.9%) | 0.38 |
| *Hypertension* | 366 (33.2%) | 188 (32.7%) | 178 (33.7%) | 0.72 |
| *Congestive Heart Failure* | 32 (2.9%) | 18 (3.1%) | 14 (2.7%) | 0.64 |
| *Diabetes Mellitus Type 2* | 221 (20.0%) | 109 (19.0%) | 112 (21.2%) | 0.35 |
| *Prior Stroke* | 124 (11.2%) | 65 (11.3%) | 59 (11.2%) | 0.95 |
| *Coronary Artery Disease* | 68 (6.2%) | 38 (6.6%) | 30 (5.7%) | 0.52 |
| *Hyperlipidemia* | 311 (28.2%) | 156 (27.1%) | 155 (29.4%) | 0.41 |

*Data are presented as median (IQR, interquartile range) for continuous variables, and n (%) for categorical variables.*
*LKW = Last Known Well; DWI = Diffusion-Weighted Imaging; ICA = Internal Carotid Artery; MCA = Middle Cerebral Artery; ACA = Anterior Cerebral Artery; PCA = Posterior Cerebral Artery.*

### Primary Analyses

In the univariable logistic sex-stratified models as shown in Table 2, greater PBV was associated with higher odds of achieving functional independence at 90-day in both male (OR=1.30; 95% CI: 1.11-1.53) and female (OR = 1.35; 95% CI: 1.13-1.61). After adjusting for DWI-FIV, age, and NIHSS, greater PBV remained independently associated with functional independence among male (aOR =1.30 per 100 cm^3^; 95% CI: 1.10-1.60), whereas this association was no longer significant among female (aOR = 1.10; 95% CI: 0.90-1.33) (Figure 1).

**Figure 1.**
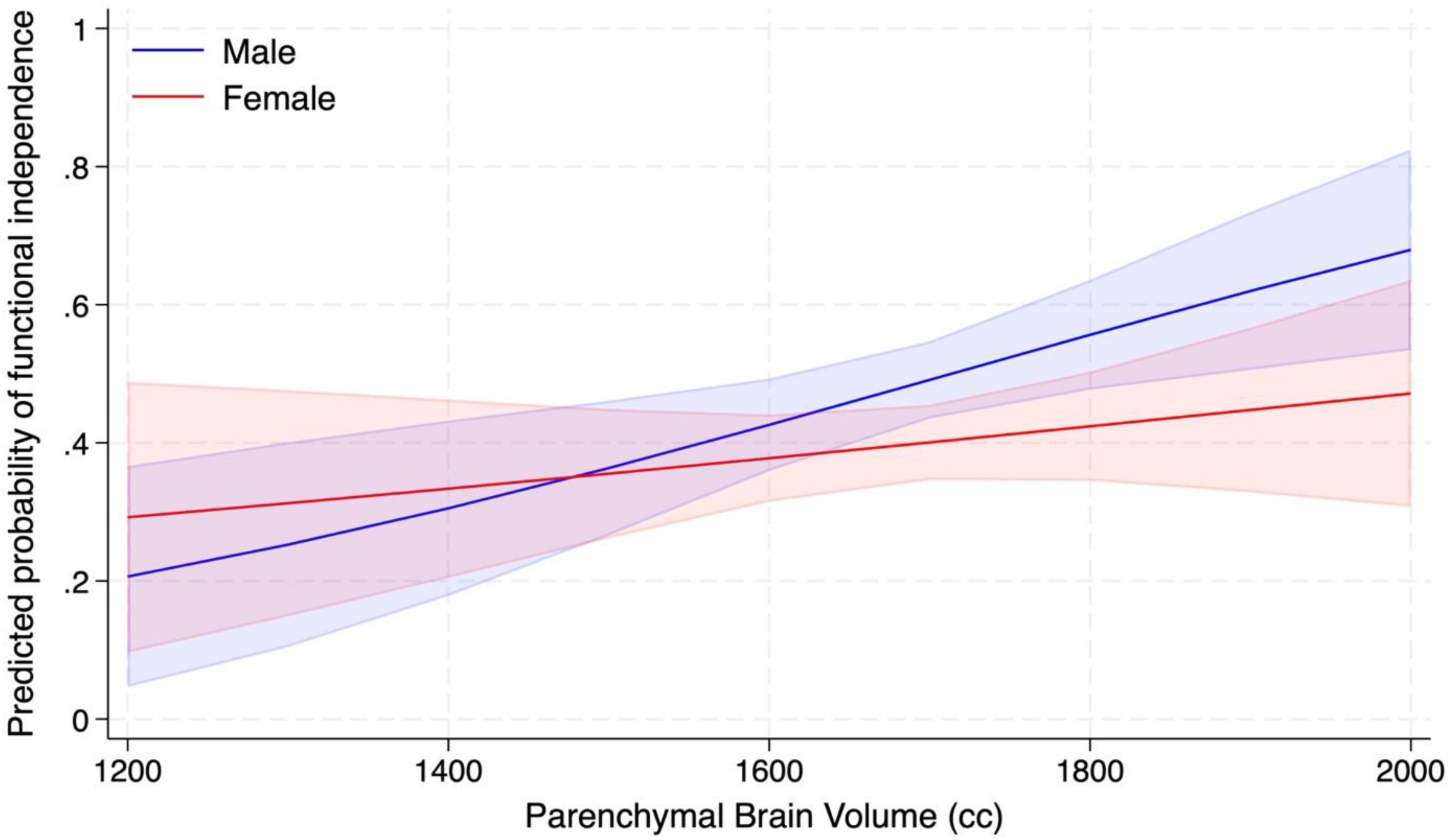
Predicted probability of functional independence across parenchymal brain volume (cm^3^) by sex. Estimates were obtained from a multivariable logistic regression model adjusted for diffusion-weighted imaging-derived final infarct volume, age, and NIHSS. The blue line represents males, and the red line represents females, with shaded areas indicating 95% confidence intervals.

**Table 2.** Associations Between Parenchymal Brain Volume (PBV) and Functional Independence (mRS 0-2).

|  |  | Univariable Analysis<br>OR (95% CI) | p-value | Multivariable Analysis<br>aOR (95% CI) | p-value |
| --- | --- | --- | --- | --- | --- |
| **Females** | PBV (per 100 cm<sup>3</sup>) | 1.35 (1.13-1.61) | 0.001 | 1.1 (0.90-1.33) | 0.328 |
|  | DWI-FIV (cm<sup>3</sup>) | 0.98 (0.97-0.99) | <0.001 | 0.98 (0.98-0.99) | <0.001 |
|  | Age (years) | 0.96 (0.95-0.98) | <0.001 | 0.99 (0.96-0.98) | <0.001 |
|  | NIHSS | 0.89 (0.87-0.92) | <0.001 | 0.92 (0.90-0.95) | <0.001 |
| **Males** | PBV (per 100 cm<sup>3</sup>) | 1.30 (1.11-1.53) | 0.001 | 1.3 (1.10-1.60) | 0.009 |
|  | DWI-FIV (cm<sup>3</sup>) | 0.98 (0.97-0.98) | <0.001 | 0.98 (0.97-0.99) | <0.001 |
|  | Age (years) | 0.98 (0.97-0.995) | 0.006 | 0.99 (0.98-1.00) | 0.225 |
|  | NIHSS | 0.90 (0.88-0.92) | <0.001 | 0.94 (0.91-0.97) | <0.001 |

Univariable and multivariable logistic regression coefficients of the association between PBV (per 100 cm<sup>3</sup>) and functional independence at 90 days. *PBV* = *Parenchymal Brain Volume*, *mRS* = *modified Rankin Scale*, *DWI-FIV* = *Diffusion-Weighted Imaging-Derived Final Infarct Volume*, *NIHSS* = *National Institute of Health Stroke Scale*.

#### Multicollinearity Assessment

Evaluation of multicollinearity demonstrated that all VIF values were below 1.5 (mean VIF = 1.32) across the predictors age, NIHSS, DWI-FIV, and PBV. Pairwise correlations among continuous predictors were modest (|r| ≤ 0.49), confirming limited shared variance and supporting the independence of covariates included in the multivariable models. These findings indicate that the attenuation of PBV’s association with functional independence among females was not attributable to multicollinearity but more likely reflects overlapping variance with age or infarct volume rather than model instability.

### Secondary Analyses

#### Sex-Stratified Associations Adjusting for Pre-Stroke mRS

To address the >20% missingness in pre-stroke mRS, we applied multiple imputation (MI, m = 160). Although the probability of missingness was unrelated to demographic or clinical variables or outcomes (p = 0.40), indicating a Missing Completely at Random (MCAR) mechanism, MI was performed to improve statistical efficiency and model robustness. In the adjusted logistic regression model using MI, where pre-stroke mRS was imputed and included with the original confounders, PBV remained non-significant among females (aOR = 1.12 per 100 cm^3^, 95% CI: 0.92–1.37; *p* = 0.26), whereas higher PBV among males (n = 575) was independently associated with greater odds of functional independence (aOR = 1.28 per 100 cm^3^, 95% CI: 1.06–1.55; *p* = 0.011). Pre-stroke mRS demonstrated a borderline inverse association with functional outcome (aOR = 0.991, 95% CI: 0.982–1.000; *p* = 0.054). The effects of infarct volume, NIHSS, and age were consistent across sexes, and adjustment for pre-stroke mRS did not alter the direction or magnitude of the associations (Supplementary Table 3).

#### Sex-Stratified Associations with 90-Day Functional Independence

For our secondary analyses, using multivariable logistic regression with 90-day functional independence as the outcome and age modeled in decades, each 10-year increase was associated with 15% lower odds of functional independence (aOR = 0.85; 95% CI: 0.72 - 0.79; *p* = 0.006) in males. Among females, the effect was more pronounced, with every 10-year increase corresponding to an 30% reduction in odds of good outcome (aOR = 0.70; 95% CI: 0.62 - 0.79; *p* < 0.001). The association between age and functional independence were significantly different between sex (*p* = 0.019), with male showing a less pronounced decline in functional independence with advancing age compared with female.

In multivariable logistic regression analysis, greater stroke severity, as measured by the NIHSS, was strongly associated with lower odds of achieving functional independence in both sexes. Each one-point increase in NIHSS reduced the probability of functional independence by 2.2% in male (95% CI: -0.0265 to -0.0183; *p* < 0.001) and by 2.4% in female (95% CI: -0.0275 to -0.0195; *p* < 0.001). The effect of stroke severity on functional independence was comparable between male and female (*p* = 0.709).

Further, in adjusted logistic regression model, DWI-FIV was inversely associated with the probability of achieving functional independence after stroke. For each cm^3^ increase in infarct volume, the probability of good outcome decreased by approximately 0.5% in men (95% CI: −0.006 to −0.004; p < 0.001) and by 0.48% in female (95% CI: −0.00611 to −0.00344; *p* < 0.001).

The change in predicted probability associated with infarct volume was nearly identical across sexes, and the interaction term was not significant (*p* = 0.779), indicating that infarct volume adversely impacts recovery to a comparable degree in males and females. No significant sex difference was observed (*p* = 0.779). (Figure 2)

**Figure 2.**
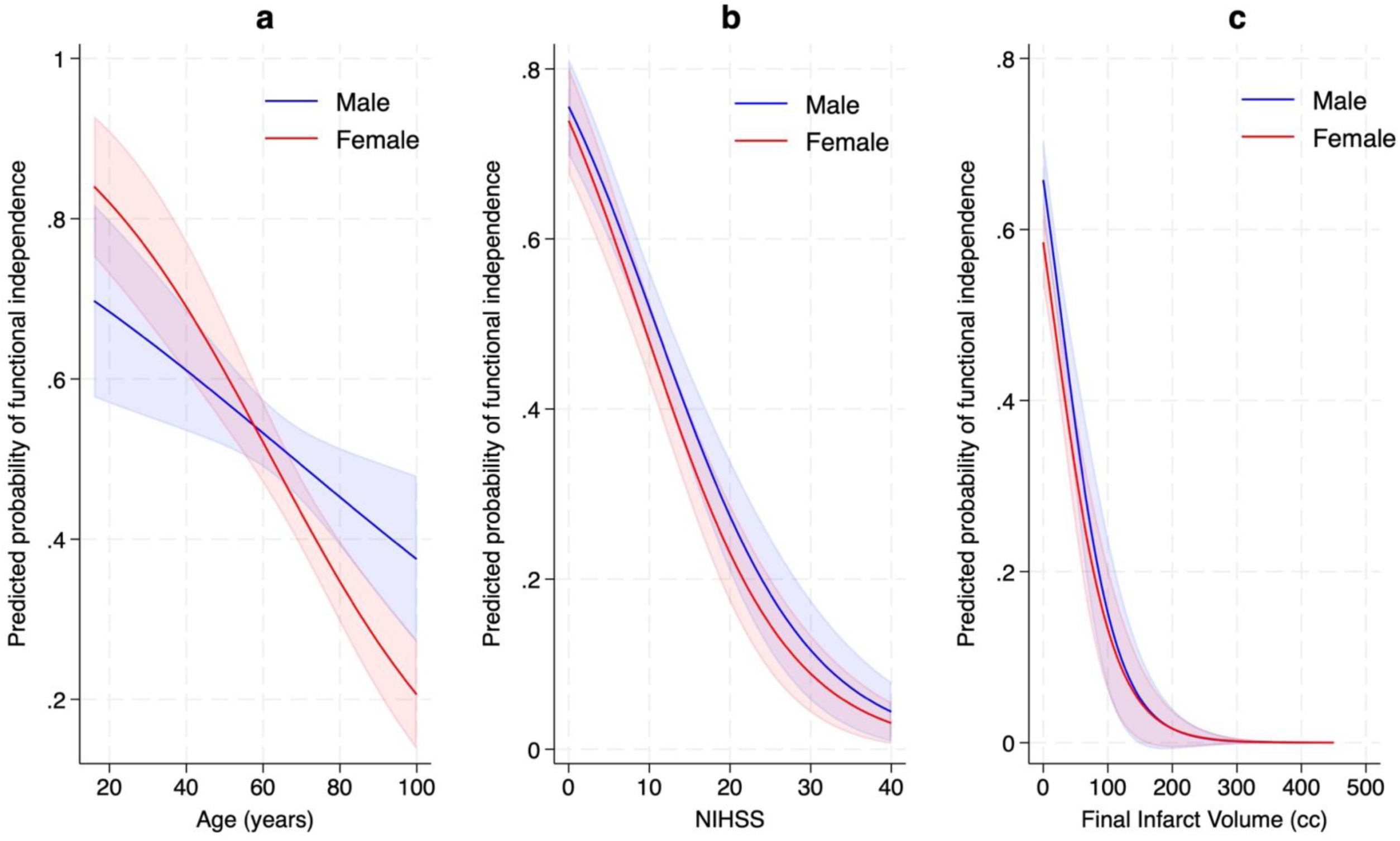
Predicted probability of functional independence across (a) age, (b) NIHSS, and (c) final infarct volume using diffused-weighted imaging by sex. Estimates were obtained from logistic regression modeling. The blue line represents males, and the red line represents females, with shaded areas indicating 95% confidence interval

#### Sex-Stratified Linear Associations Between Age and Final Infarct Volume

To further examine potential mechanisms underlying the attenuation of PBV’s effect on outcome in females, we explored the relationships among key covariates using pairwise linear regression models. Among these, a significant sex difference was identified in the association between age and DWI-FIV. Each decade of age was associated with a 4.22 cm^3^ larger DWI-FIV in females (95% CI: 0.35–8.09; *p* = 0.033), whereas no significant association was observed in males (β = –0.92 cm^3^; 95% CI: –4.75 to 2.91; *p* = 0.637). A significant age–DWI-FIV association was observed among females but not males, indicating a sex-specific difference in the relationship between infarct size and aging (Supplementary Figure S3). Additional pairwise analyses between age, NIHSS, and FIV are provided in Supplementary Figure S1–S3.

### Sensitivity Analysis

In the sensitivity analysis limited to only non-LVO patients, greater PBV remained independently associated with functional independence among male (aOR = 1.31 per 100 cm^3^; 95% CI: 1.01-1.71), whereas this association was not significant among female (aOR = 1.30; 95% CI: 0.96-1.77) after adjusting for age, NIHSS, and DWI-FIV. These results were consistent with the primary analysis, indicating similar sex-specific patterns in the relationship between PBV and functional independence at 90-days.

## Discussion

In this study of over 1100 patients with AIS, we identified variable effects of parenchymal brain volume on outcome after stroke in females versus males. Whereas larger PBV was associated with improved outcomes in males independent of other key features including age, NIHSS, and infarct volume, this relationship was not observed in females. In females, we identified that age was associated with a greater DWI-FIV, and greater NIHSS at presentation. Our finding suggest that age may abrogate some of the protective effects of PBV in females but not males.

Our results align with prior studies demonstrating that females are often older at the time of stroke. In addition, although not identified here, females in other series have greater rates of atrial fibrillation. Some studies have also found worse functional outcome in female compared to men. ^22^ ^23^ ^24^ These worse outcomes could be explained by greater severity at presentation. In the International Stroke Outcomes Study (INSTRUCT), which consisted of more than 2000 patients, found worsened outcome in female compared to males in unadjusted analysis with a non-significant difference after adjusting for presentation factors. Another study published in Japan showed worse outcome in female compared to male even after adjusting for multiple co-factors including age and NIHSS, which could suggest additional unknown factors may contribute to the prognosis after stroke in females. ^23^

DWI-FIV and PBV were comparable between males and females in our study which align with findings in the literature. ^5^ In females, age was associated with a greater DWI-FIV, which was associated with a greater NIHSS. This association may help clarify why among females age serves as the main determinant of functional outcome instead of PBV.

PBV was associated with a greater likelihood of achieving 90-day functional independence, but this effect was more pronounced in males. To ensure that this sex-specific difference was not driven by multicollinearity among covariates, variance inflation factors (VIFs) were examined for age, NIHSS, DWI-FIV, and PBV, all of which were below 1.5 (mean VIF = 1.32), indicating minimal collinearity and stable model estimates. Prior studies have shown that total brain volume decline with aging, but the rate and pattern of that decline vary between males and females with greater rates found in males. ^25^ ^26^

The mechanism by which PBV plays a diminished protective role in female relative to male is unclear. Possible explanations include hormonal changes or variable inflammation responses post stroke. Neuroinflammation plays a vital role in post-stroke injury and recovery. Neuronal damage induced by inflammation occurs both acutely and chronically following a stroke. ^27^ The sex-difference in the immune and inflammatory factors was shown to be different with age and hormonal exposure. ^28^ ^29^ Studies indicate that prolonged low gonadal steroid levels in postmenopausal females can exacerbate infarct size and inflammation, with hormone treatment showing varying effects depending on the stage of menopause. ^30^ ^31^ ^32^ Early postmenopausal females receiving hormone therapy exhibited less progression of atherosclerosis, while late postmenopausal females did not benefit. ^33^ In addition, it has been shown that ischemia-induced cell death pathways may differ in females and males. After induction of an experimental stroke in mice, females were more vulnerable to cell death induced by activation of cytochrome-C and caspase. ^30^ ^28^ This difference, along with sex-specific immune responses, may explain the age-related decline in the functional capacity of brain cells and regulation of cerebral blood flow, which may worsen neuronal damage after stroke and lead to an increased infarct size in older females.

This study has several limitations. First, although our registry was prospectively maintained, the retrospective design of this analysis introduces inherent constraints, including the potential for unmeasured confounding. Moreover, unmeasured factors, including genetic predisposition, lifestyle, post-stroke care, post-stroke depression, and hormonal data, were not included in our analysis, potentially influencing stroke outcomes and contributing to unexplained variation, particularly in females. A potential limitation of this study is the possibility of segmentation errors during PBV and DWI-FIV quantification. However, all segmentations underwent rigorous quality control to ensure accuracy and minimize this risk. Additionally, the study is enriched with thrombolysis/EVT patients due to the exclusion of those without 90-day mRS data, although in sensitivity analysis, we observed no differences in the non-LVO subset of patients.

In this large multicenter cohort, total parenchymal brain volume was robustly associated with functional independence after AIS in males but less so in females. In females, aging exerted a stronger influence on infarct size and stroke severity, suggesting that age-related vulnerability may diminish the neuroprotective effects of PBV. These findings highlight distinct sex-specific biological trajectories in cerebral aging and highlight the importance of incorporating sex and brain volume metrics into personalized stroke outcome prediction models.

## Data Availability

The data that support the findings of this study are available from the corresponding author upon reasonable request and following approval by the institutional review board and data use agreements.

## Acknowledgments

None

## Sources of Funding

Dr. Sheth reports funding from the National Institutes of Health (R01NS121154).

## Disclosures

Dr. Sheth reports grant support from the NIH (U18EB029353) as well as Viz.AI for unrelated projects. He also reports consulting fees from Viz.AI, Penumbra and Imperative Care outside of the submitted work. No other disclosures were reported.

## Supplementary Materials

### Supplementary Methods

#### Pairwise Linear Analyses

As described in the main manuscript, pairwise linear regression analyses were conducted to characterize the interrelationships between age, baseline NIHSS, and final infarct volume (FIV), stratified by sex. While the age–FIV association (presented in the main text) demonstrated a significant sex difference, the additional pairwise models—age vs. NIHSS and NIHSS vs. FIV— are provided here for completeness.

### Supplementary Results

#### Age and Stroke Severity (NIHSS)

Each decade of age was associated with an increase of 0.60 NIHSS points in men (95% CI 0.14– 1.06; *p* = 0.010) and 0.93 points in women (95% CI 0.43–1.44; *p* < 0.001), indicating a steeper age-related increase in stroke severity among females (**Supplementary Figure S1**).

#### Stroke Severity and DWI-FIV

Each one-point increase in NIHSS was associated with a 3.30 cm^3^ increase in FIV in men (95% CI 2.57–4.03; *p* < 0.001) and 3.20 cm^3^ in women (95% CI 2.48–3.92; *p* < 0.001), suggesting a comparable relationship across sexes (**Supplementary Figure S2**).

## Supplementary Table

**Supplementary Table 3.**
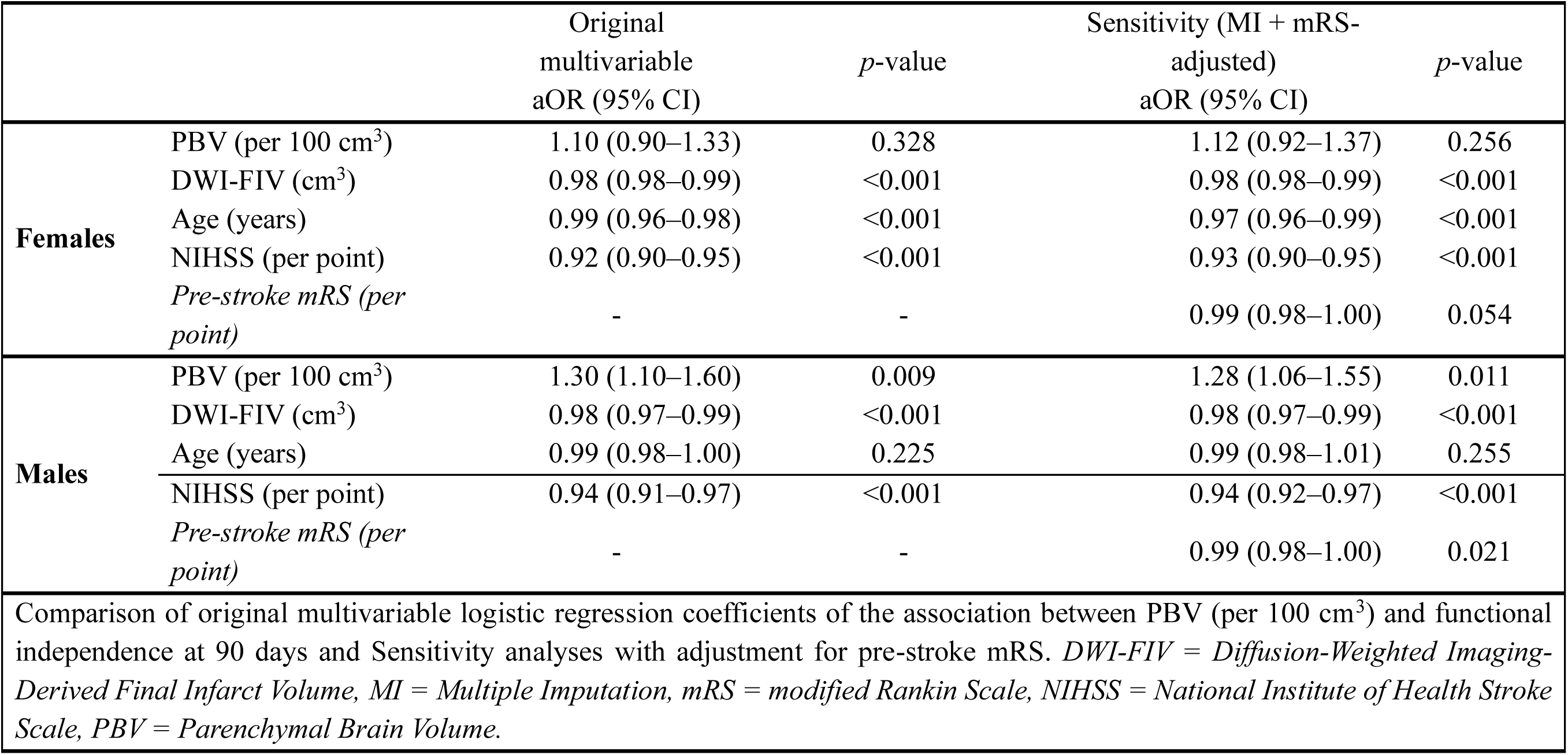
Comparison of Original Results vs. Sensitivity Analyses (mRS-Adjusted with Multiple Imputation)

## Supplementary Figures

**Supplementary Figure S1.**
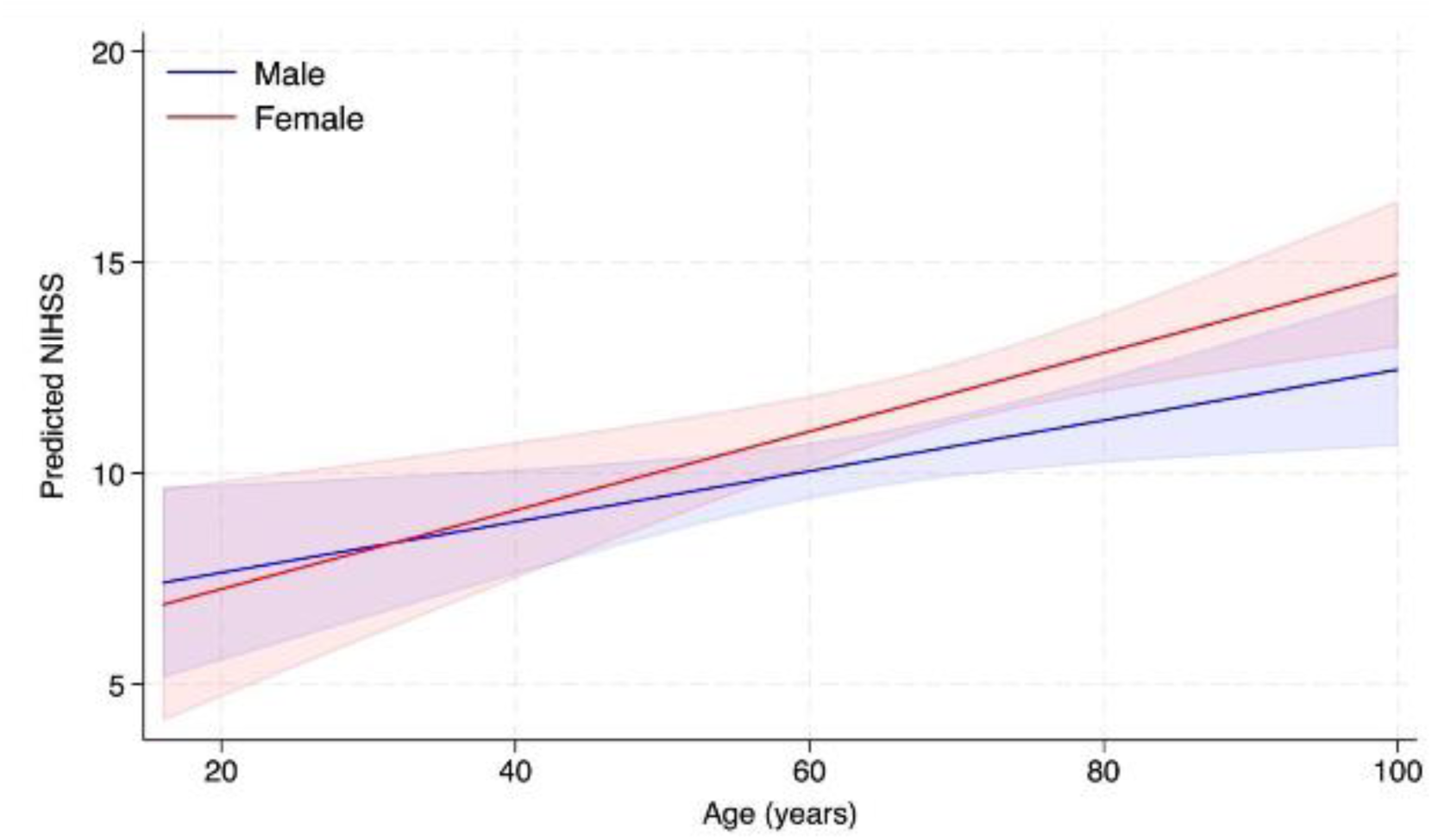
Predicted NIHSS and age by sex. Estimates were obtained from a linear regression model. The blue line represents males, and the red line represents females, with shaded areas indicating 95% confidence interval.

**Supplementary Figure S2.**
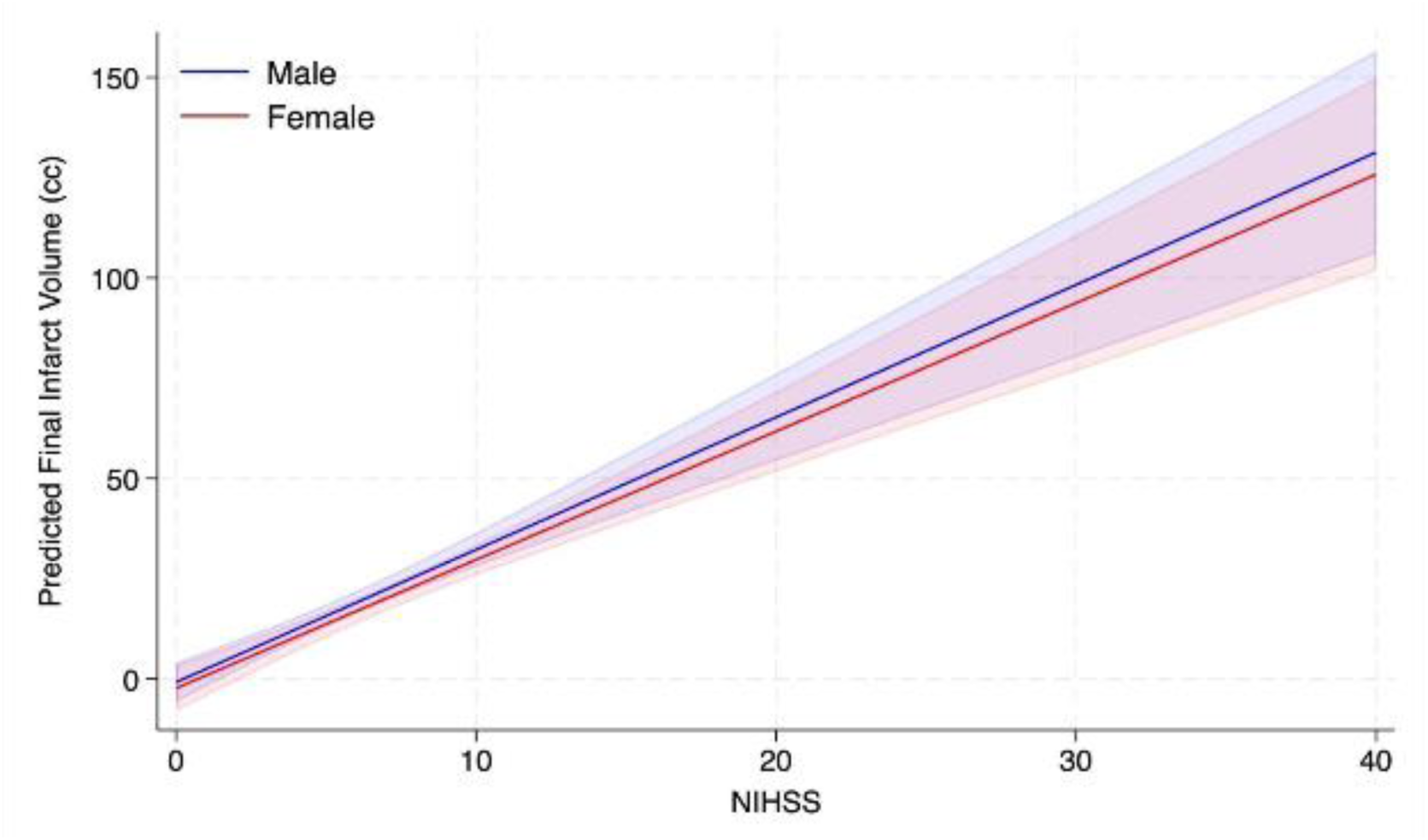
Predicted final infarct volume using diffusion-weighted imaging across NIHSS by sex. Estimates were obtained from a linear regression model. The blue line represents males, and the red line represents females, with shaded areas indicating 95% confidence interval.

**Supplementary Figure S3.**
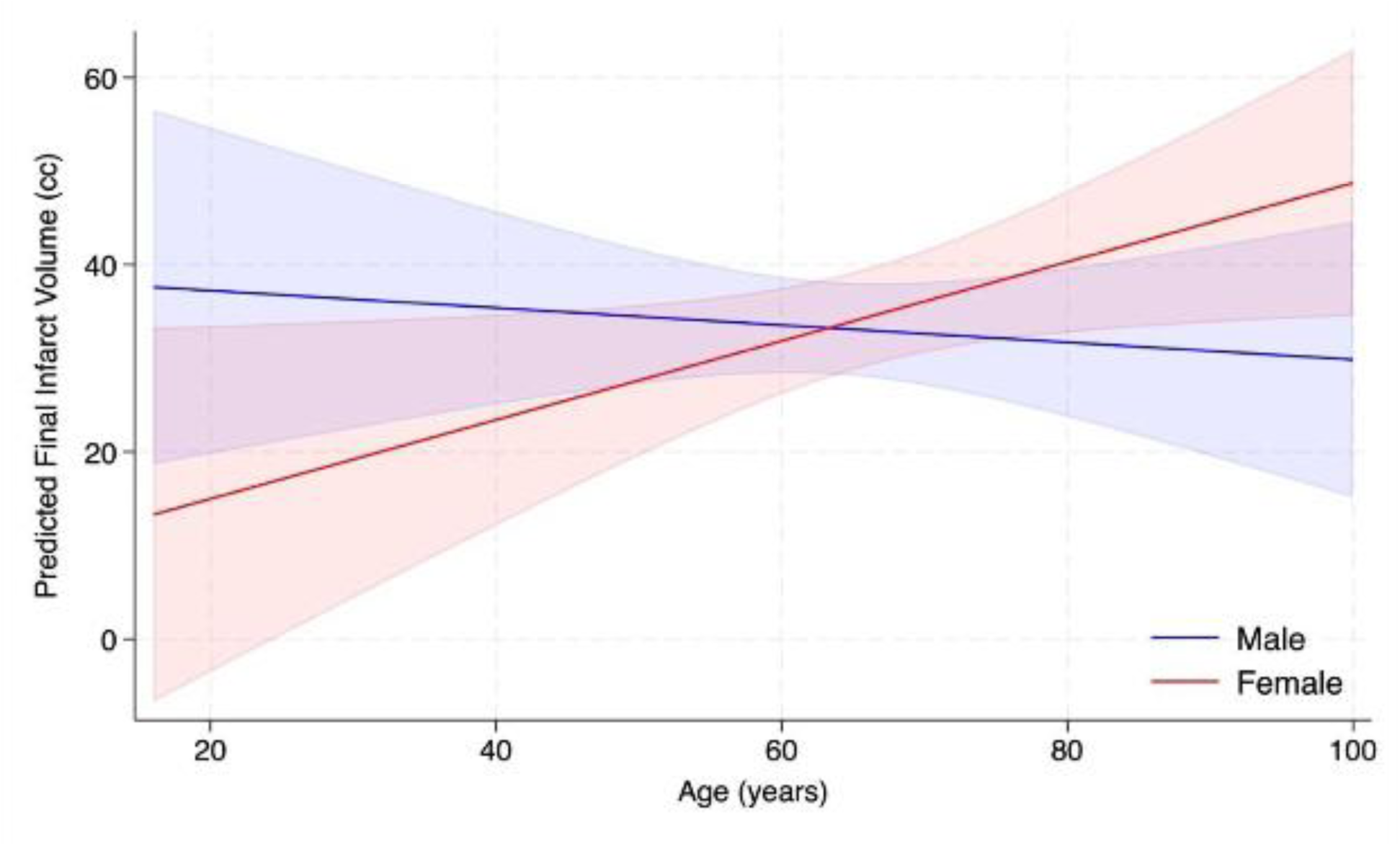
Predicted final infarct volume using diffusion-weighted imaging across age by sex. Estimates were obtained from a linear regression model. The blue line represents males, and the red line represents females, with shaded areas indicating 95% confidence interval.

## Notes

### Competing Interest Statement

The authors have declared no competing interest.

### Clinical Trial

This study is a prospective observational registry and does not meet the definition of a clinical trial requiring registration.

### Funding Statement

This study was supported by institutional resources from the Department of Neurology at UTHealth Houston. No external funding or third-party financial support was received for study design, data collection, analysis, or manuscript preparation.

### Author Declarations

This study is a prospective observational analysis using de-identified registry data. Institutional Review Board (IRB) approval was not required because no identifiable private information or patient intervention was involved, and the registry operates under institutional policies permitting use of anonymized data for research purposes. All relevant ethical guidelines were followed. The study protocol was reviewed and approved by the UTHealth Institutional Review Board.

